# A Retrospective Mixed Methods Analysis of COVID-19’s Impact on Maternal Psychosocial Health in Ghana: Lessons for Future Public Health Crises

**DOI:** 10.64898/2026.02.05.26345660

**Authors:** Ruthfirst E. A. Ayande, Gloria E. Otoo, Jessica Pearlman, Sarah Gonzalez-Nahm, Elena T. Carbone

## Abstract

**Background:** Observed impacts of the COVID-19 pandemic in countries in the global north include increasing rates of maternal anxiety and depression. However, little research has been done in sub-Saharan Africa assessing the impact of the pandemic on maternal psychosocial stress. This study aimed to examine COVID-related psychosocial impacts among Ghanaian women who were pregnant during the pandemic.

**Methods:** An online survey was administered to Ghanaian mothers with children 0-15 months old from June 2021 to January 2022. Demographic and health information, health literacy (HL) information measured by the All Aspects of Health Literacy scale (AAHL), and COVID-19 health and well-being were collected. Using the Coronavirus Anxiety Scale (CAS) and the Obsession with Coronavirus Scale (OCS), mothers were asked to recall their levels of anxiety and worry while they were pregnant during the pandemic. Generalized linear models were used to assess the association between COVID-related impacts and maternal CAS and OCS scores. In-depth interviews were conducted to gather information from participants about their experiences of pregnancy and childbirth during the COVID-19 pandemic, and thematic analyses were performed using the social support networks in health framework.

**Results:** A total of 221 mother-child dyads were analyzed. Eleven women participated in the in-depth interviews. We observed a rate of 11% for probable dysfunctional coronavirus-related anxiety and 33% for probable dysfunctional thinking about COVID-19. In adjusted models for baseline characteristics, we observed that higher HL scores corresponded with reduced CAS scores (β=-0.43, 95% CI [–0.80, –0.05] for every point increase in AAHLS scores). For COVID-related health and well-being, loss of jobs and wages predicted higher CAS scores when adjusted for HL as a main covariate (β=1.30, 95% CI [0.07, 2.53]). Participants who reported exercising during COVID had higher OCS scores compared to those who reported not exercising, when adjusting for HL (β=1.08, 95% CI [-0.01, 2.16], p=0.05). Themes emerging were categorized into three domains: COVID-19 Pandemic Impacts, COVID-19 Adaptations, and Social Support Networks.

**Conclusions and Recommendations:** This study assessing the impacts of the COVID-19 pandemic on maternal health and well-being provides valuable insights into the pandemic’s short-and medium-term effects. Given the complexity of the pandemic’s stress response, including potential intersections between psychosocial health, social support, and health literacy, public health policies must incorporate these needs into future pandemic responses. This includes leveraging telehealth and mHealth platforms for maternal mental health screening as well as providing community-based health literacy resources. Further studies examining the longer-term effects of the pandemic are also warranted.

## Introduction

At the height of the COVID-19 pandemic in 2020 through to the present day, there have been mixed results regarding the impact of COVID-19 infections on pregnancy outcomes. Some initial case reports and reviews suggested no difference in clinical findings nor increased risk of developing critical pneumonia among pregnant women with COVID-19 compared to non-pregnant women and other patients, and a low risk of vertical transmission.^1–5^ Others, however, suggested that COVID-19 infection might have caused a higher incidence of fetal distress and premature labor in pregnant women infected with COVID compared to non-COVID pregnant women.^4–6^ As more evidence becomes available, the current thinking, based largely on observations in the USA, Europe, and China, is that the virus does rarely cross the placenta.^5,7,8^ Pregnant women who present with asymptomatic COVID-19 generally do not have worse outcomes than non-COVID pregnant women,^8,9^ and the clinical course of the disease during pregnancy is about the same compared to that of non-pregnant women.^4,8^ However, in pregnant women who do develop symptoms or those with other chronic risk factors, there is a higher risk of preterm and cesarean birth.^8^ Symptomatic pregnant women, for example, are almost five times more likely to need intensive care.^8–11^ While it is still unknown if pregnancy increases the risk of infection with COVID-19, pregnancy seems to increase the risk of severe disease with COVID-19.^5,8,12^

Studies like those described above, which focus on viral infection, are crucial for assessing the biological impacts of the COVID-19 virus on pregnancy outcomes. However, it is also important to evaluate the indirect impacts the pandemic might have had on pregnant women, regardless of viral infection. These indirect indices include, but are not limited to, the psychosocial effects of the pandemic,^5,13–21^ dietary and lifestyle changes,^17,22^ as well as disruptions to maternal and child healthcare services,^17,23,24^ which inadvertently affect maternal and infant nutritional status. This is especially salient in populations in sub-Saharan Africa that reported and/or are perceived to have lower incidence rates of COVID-19 infections^25^ but might have experienced other understudied pandemic-related psycho– and socio-economic impacts.

In Western countries, increasing rates of anxiety and depression during the pandemic have been observed.^19,23,26,27^ In a cohort of 1,987 pregnant women between 18 and 48 years in Canada, Lebel and colleagues found elevated levels of anxiety and depressive symptoms in pregnant women during the pandemic, compared to similar pre-pandemic pregnancy cohorts.^26^ Other studies have also found that women from a COVID-19 cohort showed higher levels of depressive and anxiety symptoms, as compared with pre-COVID-19 women (OR = 1.94).^19^ Maternal anxiety and depression during pregnancy have been linked to poor outcomes such as preterm delivery, fetal growth retardation, increased risk of future anxiety and depression of the mother, and cognitive delays for the offspring.^27,28^ These findings, therefore, warrant investigations into the state of maternal psycho-social health during the pandemic in countries in sub-Saharan Africa where little has been done in this area and where the burden of poor birth outcomes is highest.

Pre-pandemic rates of “antenatal depression,” defined as depression that occurred during pregnancy, and measured using the Patient Health Questionnaire (PHQ) have been about 9.9% among Ghanaian pregnant women.^29^ A more recent study in 2019 of maternal postpartum depression using the Edinburgh Postnatal Depression Scale (EDPS) indicated that the prevalence of post-partum depression was 16.8% among women in Kintampo, Ghana.^30^ In Northern Ghana, Lillie and colleagues (2020) found that 19.7% of pregnant women reported symptoms of moderate to severe depression using the PHQ. Due to the varying instruments used to determine depressive symptoms across these studies, it is difficult to determine the exact pre-pandemic depressive symptoms rates in the country. However, the high rates of anxiety and depression observed in other countries during the pandemic necessitate more research, especially in low– and middle-income countries like Ghana, to assess the impact that the pandemic has had on maternal mental health, especially during the acute phase of the pandemic between 2020 and 2021.

It is important to note that the literature discussed thus far employed traditional tools used to assess pre-pandemic antenatal/postpartum psychosocial health in Ghana, which have the limitation of non-specificity regarding COVID-19. The *Coronavirus Anxiety Scale* (CAS)^32^ and the *Obsession with Coronavirus Scale* (OCS),^33^ are potentially valuable tools for identifying pandemic-specific symptoms in pregnant and postpartum populations by highlighting unique psychological stressors that emerge during and after the COVID-19 pandemic. However, they have been underexplored in pregnant and postpartum populations. These scales, developed in 2020 and validated across several populations,^34–36^ focus on anxiety, obsessive thoughts, and behaviors specifically related to the coronavirus, capturing a new layer of stress that intersects with existing postpartum challenges. While neither scale has been validated among pregnant women, they capture relevant pandemic stressors and have been used successfully among pregnant women in international settings, including India^37^, Turkey^38^ Italy,^14^ and Jamaica^18^.

The CAS measures psychosomatic symptoms of anxiety triggered by the pandemic, including physical symptoms like dizziness, paralysis, sleep disturbances, and loss of appetite,^32,39^ all of which may exacerbate depression or anxiety in mothers who are already vulnerable to emotional and physical changes. The OCS, on the other hand, specifically assesses obsessive thoughts and compulsive behaviors related to COVID-19 such as fear of infection, excessive worry about contracting the virus, and heightened health-related vigilance,^33^ which might contribute to mental strain in individuals who experience intrusive fears about their baby’s health or their own vulnerability to the virus. By isolating these pandemic-specific stress responses, both scales provide crucial insights into how the unique circumstances of the COVID-19 pandemic have created new mental health challenges for pregnant and postpartum individuals, enabling healthcare providers to better assess and address the psychological needs of this population during a time of heightened anxiety and uncertainty. In our study, we set out to retrospectively determine the prevalence of COVID-related stress and obsession among pregnant women in Ghana using an online survey. We employed a retrospective approach to ensure that participants were pregnant during the height of the pandemic, from 2020 to 2021. To our knowledge, our study is the first to use the CAS and OCS among Ghanaian women to capture COVID-related stress experiences during pregnancy.

## Methods

This was a cross-sectional, mixed-methods study to assess the impacts of the pandemic on maternal and infant health outcomes. We employed a sequential explanatory design approach for our mixed methods. Two models of interpersonal health behavior undergirded the theoretical basis of this project: the transactional model of stress and coping (TMSC), and the social network theory and analysis (SNT/A). The TMSC posits that psychological stress is a response mediated by an individual’s appraisal of internal and external environmental factors.^40^ Via cognitive appraisals (assessing an encounter as it relates to a person’s wellbeing) and coping (cognitive and behavioral efforts to adapt to the stressor), people or communities faced with a stressor assess the severity and possible impacts that the stressor might have on them, assess the resources available to them, and/or modify their behavior to reduce the impacts of the stressor. Because the COVID-19 pandemic is a stressor, we employed the TMSC in the framing of our research questions. We were interested in understanding the psychosocial outcomes of the COVID-19 pandemic via mediating factors like changes to lifestyle behaviors and maternal stress and anxiety. What was the stress response like for Ghanaian mothers during this time, and how did it impact anxiety rates? How did this stress response impact maternal health during pregnancy? What individual and environmental coping factors helped mothers become resilient during this time?

The conceptual framework of the SNT/A was employed in both the framing of our research questions and in the analysis of our qualitative results. SNT/A is based on the assumption that individuals are embedded in relations that influence their actions and identity.^41,42^ To be sure, we employed the theory of social support, and not network analysis specifically. While developing survey questionnaires and interview guides, we included questions to help us qualitatively understand our participants’ interpersonal networks in terms of types of social supports available, nature of support provided/received, and impact of the support as it pertains to coping with the birthing and parenting experience in the context of the pandemic.

Online surveys were designed and administered using REDCap v.11, a web-based data management tool hosted by the UMASS Chan Medical School.^43,44^ Survey questionnaires are available in the Supplemental Information. Prior to data collection, Institutional Review Board (IRB) approval was obtained from the University of Massachusetts Amherst in accordance with the principles outlined in the Belmont Report. Additionally, the study adhered to the guiding principles of the Declaration of Helsinki. All participants completed and signed a consent form prior to enrollment in the study. Details of the study population and measurement instruments are described below.

## Study Participants

The study was conducted among women with children between 0 and 15 months old who had access to the internet (for participants who could self-administer the survey) or a phone (for interviewer-administered surveys) and were willing to complete the survey. Recruitment lasted from July 2021 to January 2022. Of note, from July 2021 to November 2021, the age criterion for children was 0-12 months, however, this was extended to 15 months in December 2021 through January 2022 because recruitment took longer than anticipated. Women were recruited through social media platforms (Facebook, WhatsApp, Instagram) and by word-of-mouth for those who were not on social media or did not have access to the internet. To limit selection bias as much as possible, research assistants based in three major regions of Ghana facilitated phone calls to mothers who lacked internet access. Notably, we were aware that online only recruitment would likely bias our participants toward internet users who may be more literate and/or have higher socioeconomic status. We therefore employed these three research assistants, one in the Northern Region of the country, one in the Ashanti Region (middle belt of the country) and the other in the Greater Accra Region (Southern belt of the country), to facilitate on-the-ground community outreach, including contacting baby weighing centers and liaising with health professionals to identify eligible participants. All research assistants held nutrition-related degrees and completed IRB training to meet standards set by the University of Massachusetts.

Women with children older than the specified age cutoff were excluded from the study to minimize the risk of recall bias and ensure that pregnancies occurred during the peak period of the COVID-19 pandemic, between 2020 and 2021. Women who reported a previous diagnosis of postpartum depression were excluded. Participants were also excluded if they were non-Ghanaian or were currently pregnant (pregnancy not being carried at the peak of the pandemic). There were no other restrictions besides those described above.

Flyers and advertisements were circulated on Facebook, WhatsApp, and other social media platforms, as well as on the project website, targeting women who met the study’s inclusion criteria. Women were provided with a written project summary and directed to a link to assess eligibility and enroll on the study. Once women met the screening criteria, they proceeded to the consent form. Women had to sign the consent form before proceeding to the main study. Women who were called on the phone and could not access the project online provided verbal consent after interviewers explained the details of the project as outlined on the consent form to them. The project was in two phases. Phase 1 involved the online/phone survey, and phase 2 was concerned with a nutrition and health education intervention to improve maternal and infant dietary diversity. The consent form covered all aspects of the study in phases 1 and 2, including both quantitative and qualitative components (i.e., participants did not sign separate consent forms for each aspect of the study). The focus of this manuscript is on phase 1 of the project. For the qualitative aspects of the study, a conveniently selected subsample of eleven women who had completed the baseline and Phase 1 questionnaires and indicated their prior interest were invited to participate in in-depth interviews during the project’s second phase. The only criteria for participation were enrollment in Phase 2 of the study and willingness to participate in the interview. Interviews were conducted using a semi-structured guide (see Supplemental data 7), and were offered by phone, via Zoom, WhatsApp, or Facebook Messenger, based on the woman’s preference.

Mothers were provided with a cash incentive to cover their data costs. The breakdown is as follows: $10 for phase 1 mothers (exchange rate at the time was GHC 50); $10 for intervention phase 2 (GHC 100); and $5 for in-depth interviews during phase 2 (GHC 50).

## Screening, Demographic and Health Information

At baseline, women completed a screening questionnaire used to determine eligibility for the study. The first page of the survey was the screening questionnaire, and the second page was the baseline questionnaire (socio-demographic survey and provided health information). The socio-demographic information collected included maternal age, occupation, ethnicity, socioeconomic status, and education. Participants provided the following information from their medical records: date of enrollment at an antenatal clinic/hospital, number of antenatal sessions completed, diagnosis of gestational diabetes, and diagnosis of gestational hypertension. Participants could progress through the survey only after meeting eligibility criteria (the REDCap survey was designed to automatically end surveys for participants who did not meet any of the criteria).

## COVID-related Stress and Anxiety

Two questionnaires were administered online to examine maternal COVID-19-related anxiety and stress during pregnancy. The Coronavirus Anxiety Scale (CAS)^32^ was used to assess maternal COVID-19-related anxiety. This 5-item scale examines dysfunctional anxiety symptoms associated with the coronavirus pandemic. The Obsession with COVID-19 Scale (OCS)^33^ is a 4-item measure that assesses persistent and disturbed thinking about COVID-19. Both tools have been validated; the CAS-score has a Reliability (αs) > .90 whereas the OCS-score has a Reliability (αs) > .83.^32,33^

## COVID Health and Wellbeing Questionnaire

We adapted the COVID-19 Impact on Health and Well-being Survey developed by Robledo in 2020 and available in the REDCap instrument Library, to assess maternal COVID-19 health and wellbeing.^45^ To limit participant burden, we abridged the questionnaire and only asked questions relevant to maternal experiences of shelter-in-place/stay-at-home directives, impact on livelihood, and impact on lifestyle changes (e.g., physical activity/exercise).

## Data Quality Checks

To ensure the integrity of the online survey data and confirm that survey respondents were individuals who truly met the inclusion criteria, the research team conducted follow-up calls to participants who completed phase 1 to verify the information they provided on the surveys. Participants who could not validate the information provided on the surveys, especially relevant information pertaining to themselves and their infant’s medical and birth records, were removed from the analytical dataset.

## Qualitative Interviews

Eleven (11) women from the subsample of participants who indicated availability for the second phase of the project, expressed interest in participating in the interviews, and responded to follow-up requests, were invited to take part in an in-depth interview session. Interviews ranged from 40 minutes to an hour. A semi-structured interview guide was developed and used for the interview (See Supplemental Information 7).

Women were asked questions about their pregnancy experiences during the COVID-19 pandemic, knowledge of the COVID-19 pandemic, how worried they felt about their health and the health of their child during the pandemic, access to maternal and childcare centers during pregnancy, and sources of social support available to them.

## Statistical Analysis

We conducted univariate analyses on demographic data to describe the distribution of COVID-19-related stress and anxiety and all demographic variables. We calculated the percent distribution for categorical variables (maternal educational status, parity, probable dysfunctional coronavirus anxiety, and probably dysfunctional coronavirus obsession), and means were computed for continuous variables (maternal age, AAHLS score, total CAS, and OCS scores). We evaluated the unadjusted and adjusted relationship between maternal demographic characteristics (age, parity, education, AAHL individual domain and total scores) and COVID-19-related anxiety and COVID-related obsession to determine which demographic variables were associated with each outcome.

For our main project aim, assessing the COVID-19 pandemic impacts on maternal psychosocial stress, we built generalized linear models to assess the association between COVID-related health impacts (predictors) and maternal CAS and OCS scores. Our predictors were COVID-19 infection while pregnant, COVID-sanctioned restrictions, Loss of job/wages and changes to family financial status, physical activity, and changes to exercise habits during COVID. Due to the small number of respondents who reported COVID-19 infection during pregnancy and some not knowing their status during pregnancy, as well as a small number reporting not being under current COVID-19 lockdowns, we only focused on past COVID-19 lockdowns, loss of wages due to COVID-19, working in-person, the household financial situation, and exercise. After assessing the unadjusted relationship between these variables and each of our outcomes, we built three adjusted models for each COVID-impact exposure. Model 1 adjusted for AAHLS total score because HL was a primary covariate, Model 2 adjusted for AAHLS total score and maternal education, and model 3 adjusted for AAHLS total score and household financial situation. The adjustments in models 2 and 3 were made to account for additional socioeconomic context. To ensure that our models met the assumptions for regression analysis, we performed diagnostic tests for the presence of collinearity and heteroscedasticity, of which none of these were violated. Per the central limit theorem, we did not include normality of errors as a required assumption in our models because our sample size was over 200.

We performed all our data cleaning and analysis using SAS 9.4 (SAS Institute Inc, Cary NC).

## Qualitative Data Analysis

Analysis of qualitative data was managed using NVivo version 12 Plus (QSR, 2018). Audio recordings of the in-depth interviews were transcribed verbatim and exported into NVivo where they were coded using open and axial coding and then thematized to identify emergent themes.^46^ The social support networks and social support to health theory served as the framework of our thematic analysis.^41^ Themes emerging from the interviews were used to deepen understanding of the quantitative data and are presented before the discussion section of this paper.

## Results

### Characteristics of Study Participants

After accounting for incomplete consent forms and surveys, 291 surveys initially remained prior to additional data quality checks (Figure 1). Following data quality checks via follow-up phone calls and data queries, 70 records (24%) were excluded, of which 21.6% were due to unverifiable or invalid medical records, 2.1% evaded form logic (i.e., participants were not truly eligible), and 0.3% resulted from a duplicate survey (1 record).

**Figure 1:**
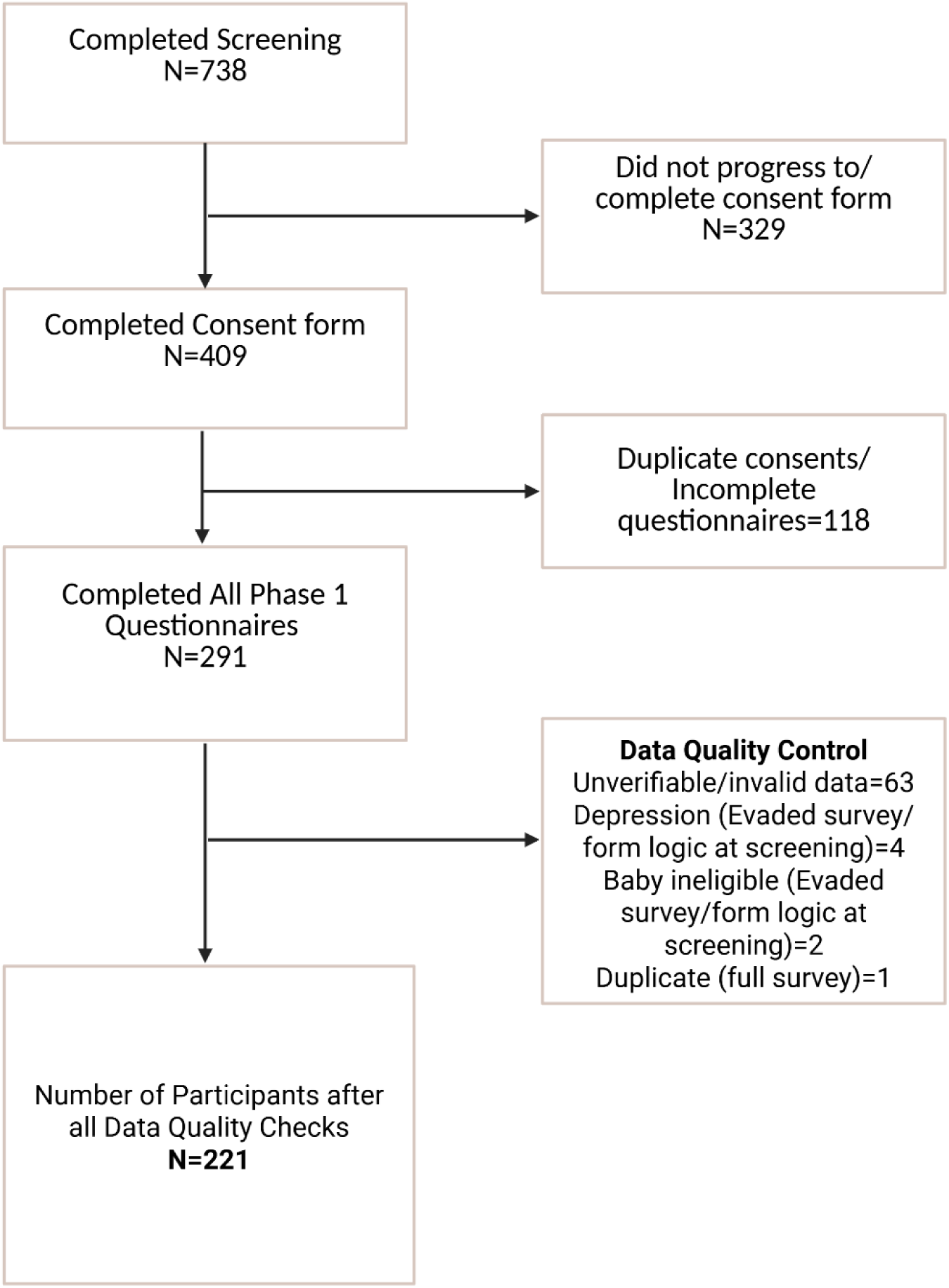
Flow diagram of Project Showing Number of Participants from Screening through Enrollment and Completion of Questionnaires. Created with BioRender.com.

A final total of 221 Ghanaian women remained, following data quality checks (See Table 1 and Figure 1). The mean age of the mothers in the sample was 29.5 ± 5.0 years. Forty-four percent (44%) of the mothers were primiparous (i.e., index infant was their first child), and 33% had a secondary school education or less, with the majority of the mothers (67%) having either a tertiary or postgraduate education. Based on a total possible score of 3 each, the average functional health literacy, critical health literacy, and communicative health literacy scores were 2.5±0.7, 2.4±0.6 and 2.7±0.5, respectively. When aggregated into an overall AAHLS total score, the mean was 7.5±1.5 (out of a total possible score of 9). (Table 1).

**Table 1:** Distribution of sociodemographic and health characteristics of the study sample.

| Characteristic | Distribution<br>(N=221) |
| --- | --- |
| **Maternal Age (years)** | 29.5±5.0<sup>1</sup> |
| **Primiparous** |  |
| No | 123 (56)<sup>2</sup> |
| Yes | 98 (44) |
| **Maternal Educational Status** |  |
| Secondary School or less | 73 (33) |
| Tertiary or Postgraduate School | 148 (67) |
| **All Aspect of Health Literacy Scale** |  |
| Functional Health Literacy | 2.5 ± 0.7 |
| Critical Health Literacy | 2.4± 0.6 |
| Communicative Health Literacy | 2.7 ± 0.5 |
| AAHLS total score | 7.5± 1.5 |
| **COVID-related Anxiety and Obsession** |  |
| CAS score (continuous) | 3.5 ± 3.5 |
| **CAS categorized** |  |
| Less than 9 | 196 (89) |
| Greater than 9 | 25 (11) |
| OCS score (continuous) | 5.4±3.6 |
| **OCS categorized** |  |
| Less than 7 | 148 (67) |
| Greater than 7 | 73 (33) |

<sup>1</sup>Mean±SD, <sup>2</sup>N(%). CAS: Coronavirus Anxiety Scale. OCS: Obsession with Coronavirus Scale.

### Prevalence of Coronavirus-related Stress and Coronavirus-related Obsession

The mean CAS score among the sample population was 3.5±3.5 (out of a total possible score of 20), and the mean OCS score among the sample population was 5.4±3.6 (out of a total possible score of 16). Based on the suggested CAS cutoff of 9,^32^ the proportion of all participants with probable dysfunctional coronavirus-related anxiety was 11%. Based on the suggested OCS cutoff of 7,^33^ the proportion of all participants with probable dysfunctional thinking about COVID-19 was 33% (Table 1).

### COVID-related Impacts on Health and Wellbeing

Table 2 shows results for the COVID-related health and well-being of mothers. A large proportion of participants (80%) indicated that they did not get infected with COVID while pregnant. Twenty percent of participants (20%) either reported that they got infected with COVID (2%) or did not know if they got COVID while pregnant (18%). Forty-eight percent (48%) of participants reported experiencing a past lockdown; however, almost all participants (98%) reported not being currently under lockdown. Fewer than a fifth of participants (16%) reported that they lost their job or wages because of COVID, and 57% reported they were working in person. When asked about their current financial situation at home, more than a third (35%) reported having to cut back, and 16% reported that they could not make ends meet.

**Table 2:** COVID-related impacts on health and well-being.

| COVID Impact | Distribution<br>(N=221) |
| --- | --- |
|  | N(%) |
| COVID-19 infection while pregnant |  |
| No | 176 (80) |
| Yes | 4 (2) |
| Don't know | 40 (18) |
| COVID-sanctioned Restrictions |  |
| *Lockdown in the past* |  |
| No | 114 (52) |
| Yes | 104 (48) |
| *Currently lockdown* |  |
| No | 215 (98) |
| Yes/Don't know | 4 (2) |
| Loss of job or wages because of COVID-19 |  |
| No | 185 (84) |
| Yes | 34 (16) |
| Working in-person |  |
| No | 94 (43) |
| Yes | 124 (57) |
| Financial Situation at Home during COVID-19 |  |
| Comfortable, with extra | 18 (8) |
| Enough, but not extra | 90 (41) |
| Have had to cut back | 77 (35) |
| Cannot make ends meet | 34 (16) |
| Exercise during COVID (N=220) |  |
| No exercise | 74 (34) |
| Exercise | 146 (66) |
| Change in Exercise (N=142) |  |
| Less | 80 (56) |
| Same as before pandemic | 27 (19) |
| More than before pandemic | 35 (25) |

One hundred and forty-six participants, representing 66% of respondents, reported exercising during the pandemic. Among those who exercised, 56% reported that they exercised less than they did before the pandemic, while 44% reported that they exercised the same (19%) or more (25%) than they did before the pandemic (Table 2).

### Relationship between Maternal Demographic Characteristics and COVID Anxiety, and Obsession with COVID

In our bivariate analysis, we observed that maternal educational status and the individual domains of health literacy were all associated with lower CAS scores. Mothers with a tertiary education or higher scored 1.96 points lower (β=-1.96, 95% CI [–2.90, –1.02]) on the CAS scale compared with mothers with secondary or lower education. The relationship was attenuated and not significant when the model was adjusted for maternal age, primiparity, and AAHLS total score (β = –1.12, 95% CI [-2.39, 0.15]) (Table 3).

**Table 3:**
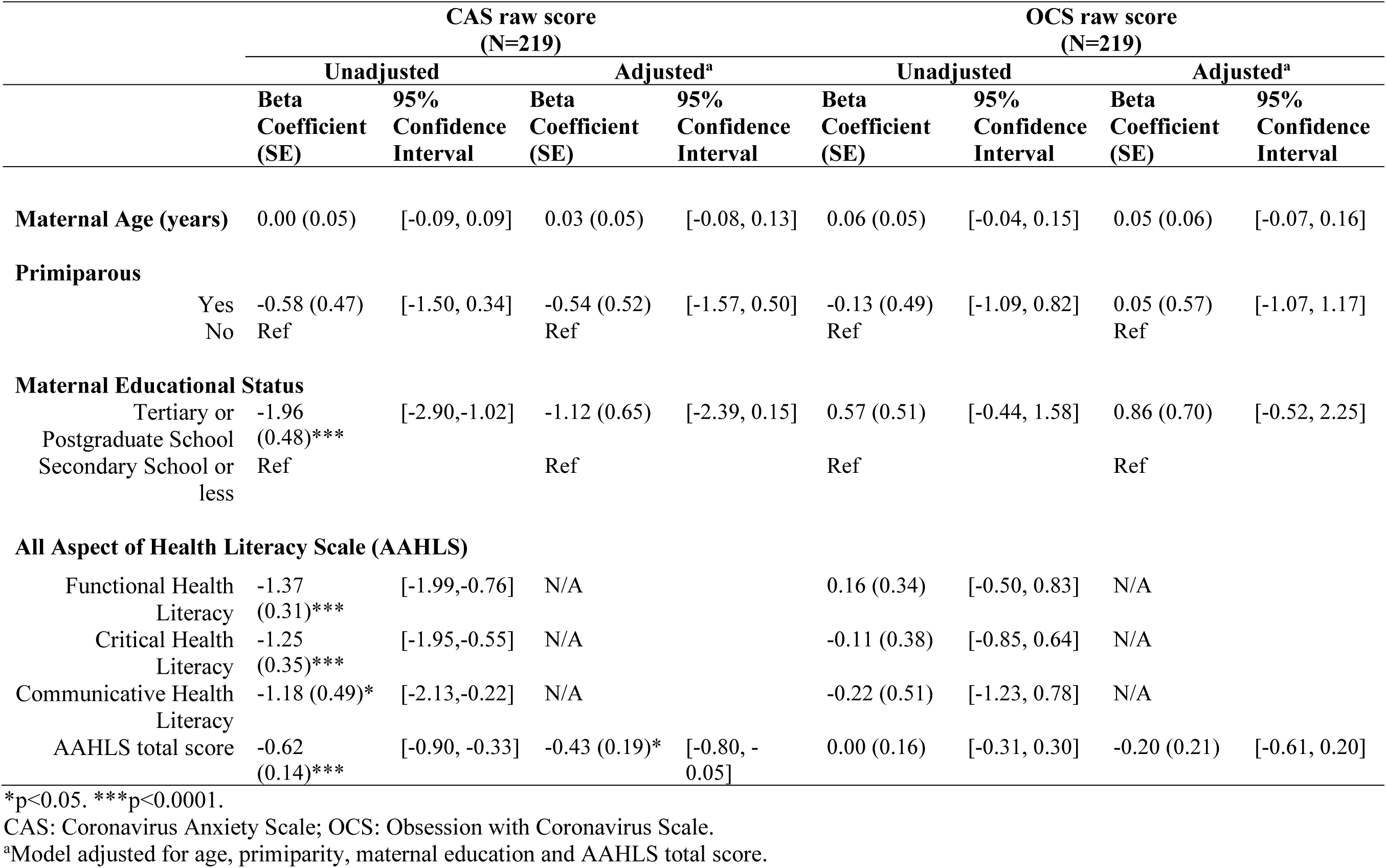
Relationship between maternal baseline characteristics and CAS and OCS scores.

|  | CAS raw score<br>(N=219) |  |  |  | OCS raw score<br>(N=219) |  |  |  |
| --- | --- | --- | --- | --- | --- | --- | --- | --- |
|  | Unadjusted |  | Adjusted<sup>a</sup> |  | Unadjusted |  | Adjusted<sup>a</sup> |  |
|  | Beta<br>Coefficient<br>(SE) | 95%<br>Confidence<br>Interval | Beta<br>Coefficient<br>(SE) | 95%<br>Confidence<br>Interval | Beta<br>Coefficient<br>(SE) | 95%<br>Confidence<br>Interval | Beta<br>Coefficient<br>(SE) | 95%<br>Confidence<br>Interval |
| **Maternal Age (years)** | 0.00 (0.05) | [-0.09, 0.09] | 0.03 (0.05) | [-0.08, 0.13] | 0.06 (0.05) | [-0.04, 0.15] | 0.05 (0.06) | [-0.07, 0.16] |
| **Primiparous** |  |  |  |  |  |  |  |  |
| Yes | -0.58 (0.47) | [-1.50, 0.34] | -0.54 (0.52) | [-1.57, 0.50] | -0.13 (0.49) | [-1.09, 0.82] | 0.05 (0.57) | [-1.07, 1.17] |
| No | Ref |  | Ref |  | Ref |  | Ref |  |
| **Maternal Educational Status** |  |  |  |  |  |  |  |  |
| Tertiary or Postgraduate School | -1.96 (0.48)*** | [-2.90,-1.02] | -1.12 (0.65) | [-2.39, 0.15] | 0.57 (0.51) | [-0.44, 1.58] | 0.86 (0.70) | [-0.52, 2.25] |
| Secondary School or less | Ref |  | Ref |  | Ref |  | Ref |  |
| **All Aspect of Health Literacy Scale (AAHLS)** |  |  |  |  |  |  |  |  |
| Functional Health Literacy | -1.37 (0.31)*** | [-1.99,-0.76] | N/A |  | 0.16 (0.34) | [-0.50, 0.83] | N/A |  |
| Critical Health Literacy | -1.25 (0.35)*** | [-1.95,-0.55] | N/A |  | -0.11 (0.38) | [-0.85, 0.64] | N/A |  |
| Communicative Health Literacy | -1.18 (0.49)* | [-2.13,-0.22] | N/A |  | -0.22 (0.51) | [-1.23, 0.78] | N/A |  |
| AAHLS total score | -0.62 (0.14)*** | [-0.90, -0.33] | -0.43 (0.19)* | [-0.80, -0.05] | 0.00 (0.16) | [-0.31, 0.30] | -0.20 (0.21) | [-0.61, 0.20] |

\*p<0.05. \*\*\*p<0.0001.
CAS: Coronavirus Anxiety Scale; OCS: Obsession with Coronavirus Scale.
<sup>a</sup>Model adjusted for age, primiparity, maternal education and AAHLS total score.

For every point increase in functional health literacy, critical health literacy and communicative health literacy, mothers significantly scored lower on the CAS scale by 1.37 (β=-1.37, 95% CI [-1.99, –0.76]), 1.25 (β=-1.35, 95% CI [–1.95, –0.55]), and 1.18 (β=-1.18, 95% CI [–2.13, –0.22]) points respectively. For every one-point increase in maternal aggregated AAHLS scores, we observed a 0.62-point decrease (β = – 0.62, 95% CI = [–0.90, –0.33]) in CAS score. This relationship persisted in our model adjusting for maternal age, parity, and maternal education (β=-0.43, 95% CI [-0.80,-0.05]) (Table 3), making health literacy a stronger independent predictor of coronavirus anxiety compared to education, and therefore our primary covariate of interest.

We did not find statistically significant associations between any of our demographic information and OCS scores in both our unadjusted and adjusted models (Table 3).

### Factors Associated with COVID Anxiety and Obsession with COVID

In our unadjusted models, we observed no statistically significant associations between past lockdown, loss of wages, and working in person on CAS (Table 4) and OCS scores (Table 5)

**Table 4:**
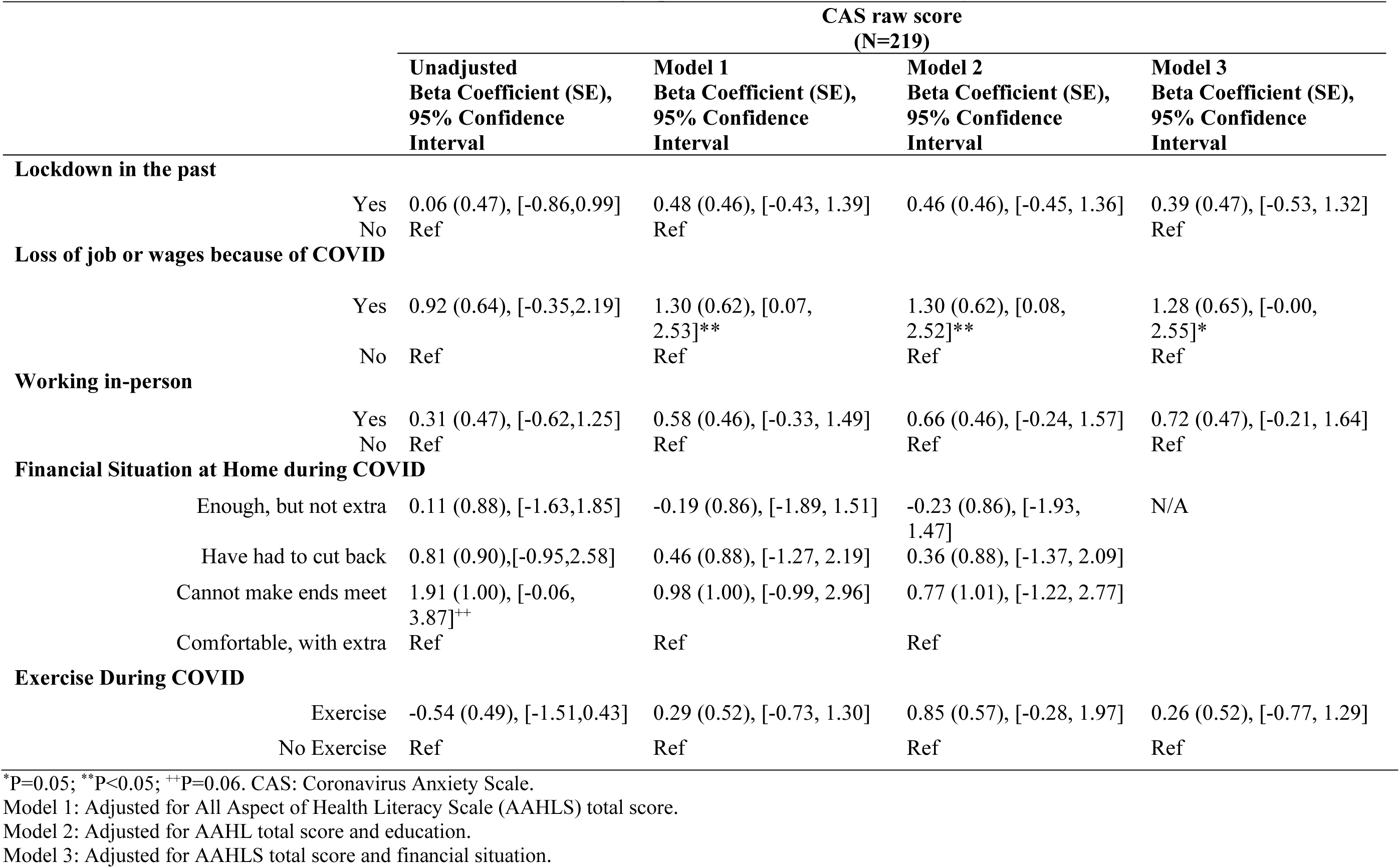
Association between COVID-related health and well-being impacts and COVID-related Anxiety Scores.

|  |  | CAS raw score<br>(N=219) |  |  |  |
| --- | --- | --- | --- | --- | --- |
|  |  | Unadjusted<br>Beta Coefficient (SE),<br>95% Confidence<br>Interval | Model 1<br>Beta Coefficient (SE),<br>95% Confidence<br>Interval | Model 2<br>Beta Coefficient (SE),<br>95% Confidence<br>Interval | Model 3<br>Beta Coefficient (SE),<br>95% Confidence<br>Interval |
| **Lockdown in the past** |  |  |  |  |  |
|  | Yes | 0.06 (0.47), [-0.86,0.99] | 0.48 (0.46), [-0.43, 1.39] | 0.46 (0.46), [-0.45, 1.36] | 0.39 (0.47), [-0.53, 1.32] |
|  | No | Ref | Ref |  | Ref |
| **Loss of job or wages because of COVID** |  |  |  |  |  |
|  | Yes | 0.92 (0.64), [-0.35,2.19] | 1.30 (0.62), [0.07, 2.53]** | 1.30 (0.62), [0.08, 2.52]** | 1.28 (0.65), [-0.00, 2.55]* |
|  | No | Ref | Ref | Ref | Ref |
| **Working in-person** |  |  |  |  |  |
|  | Yes | 0.31 (0.47), [-0.62,1.25] | 0.58 (0.46), [-0.33, 1.49] | 0.66 (0.46), [-0.24, 1.57] | 0.72 (0.47), [-0.21, 1.64] |
|  | No | Ref | Ref | Ref | Ref |
| **Financial Situation at Home during COVID** |  |  |  |  |  |
|  | Enough, but not extra | 0.11 (0.88), [-1.63,1.85] | -0.19 (0.86), [-1.89, 1.51] | -0.23 (0.86), [-1.93, 1.47] | N/A |
|  | Have had to cut back | 0.81 (0.90),[-0.95,2.58] | 0.46 (0.88), [-1.27, 2.19] | 0.36 (0.88), [-1.37, 2.09] |  |
|  | Cannot make ends meet | 1.91 (1.00), [-0.06, 3.87]<sup>++</sup> | 0.98 (1.00), [-0.99, 2.96] | 0.77 (1.01), [-1.22, 2.77] |  |
|  | Comfortable, with extra | Ref | Ref | Ref |  |
| **Exercise During COVID** |  |  |  |  |  |
|  | Exercise | -0.54 (0.49), [-1.51,0.43] | 0.29 (0.52), [-0.73, 1.30] | 0.85 (0.57), [-0.28, 1.97] | 0.26 (0.52), [-0.77, 1.29] |
|  | No Exercise | Ref | Ref | Ref | Ref |

\*P=0.05; \*\*P<0.05; ++P=0.06. CAS: Coronavirus Anxiety Scale.
Model 1: Adjusted for All Aspect of Health Literacy Scale (AAHLS) total score.
Model 2: Adjusted for AAHL total score and education.
Model 3: Adjusted for AAHLS total score and financial situation.

**Table 5:** Association between COVID-related health and well-being impacts and Obsession with Coronavirus Scale Scores.

|  |  | OCS raw score<br>(N=219) |  |  |  |
| --- | --- | --- | --- | --- | --- |
|  |  | Unadjusted<br>Beta Coefficient (SE),<br>95% Confidence<br>Interval | Model 1<br>Beta Coefficient (SE),<br>95% Confidence<br>Interval | Model 2<br>Beta Coefficient (SE),<br>95% Confidence<br>Interval | Model 3<br>Beta Coefficient (SE),<br>95% Confidence<br>Interval |
| **Lockdown in the past** |  |  |  |  |  |
|  | Yes | 0.66 (0.49), [-0.29, 1.62] | 0.70 (0.50), [-0.28, 1.68] | 0.72 (0.50), [-0.26, 1.70] | 0.65 (0.50), [-0.33, 1.64] |
|  | No | Ref | Ref | Ref | Ref |
| **Loss of job or wages because of COVID** |  |  |  |  |  |
|  | Yes | 0.38 (0.67), [-0.94, 1.70] | 0.39 (0.68), [-0.95, 1.73] | 0.39 (0.68), [-0.94, 1.73] | 0.35 (0.69), [-1.02, 1.71] |
|  | No | Ref | Ref | Ref | Ref |
| **Working in-person** |  |  |  |  |  |
|  | Yes | 0.13 (0.49), [-0.85, 1.10] | 0.13 (0.50), [-0.86, 1.11] | 0.06 (0.50), [-0.93, 1.04] | 0.28 (0.50), [-0.71, 1.27] |
|  | No | Ref | Ref | Ref | Ref |
| **Financial Situation at Home during COVID** |  |  |  |  |  |
|  | Enough, but not extra | 0.88 (0.91), [-0.92, 2.68] | 0.91 (0.92), [-0.90, 2.72] | 0.96 (0.91), [-0.84, 2.76] | N/A |
|  | Have had to cut back | 1.46 (0.93), [-0.36, 3.29] | 1.50 (0.93), [-0.34, 3.38] | 1.63 (0.93), [-0.20, 3.47] |  |
|  | Cannot make ends meet | 1.66 (1.03), [-0.37, 3.69] | 1.76 (1.07), [-0.35, 3.86] | 2.04 (1.07), [-0.08, 4.15]<sup>++</sup> |  |
|  | Comfortable, with extra | Ref | Ref | Ref |  |
| **Exercise During COVID** |  |  |  |  |  |
|  | Exercise | 0.92 (0.51)[-0.08, 1.91] | 1.08 (0.55), [-0.01, 2.16]* | 0.87 (0.61), [-0.34, 2.08] | 1.02 (0.55), [-0.06, 2.11]<sup>++</sup> |
|  | No Exercise | Ref | Ref | Ref | Ref |

\*P=0.05; ++P=0.06. OCS: Obsession with Coronavirus Scale
Model 1: Adjusted for All Aspect of Health Literacy Scale (AAHLS) total score.
Model 2: Adjusted for AAHL total score and education.
Model 2: Adjusted for AAHLS total score and financial situation.

For the financial situation at home predictor, we observed trending results where participants who reported that they could not make ends meet had a 1.91 higher CAS scores (β=1.91, 95% CI [-0.06, 3.87], p=0.06) compared to participants who reported they were comfortable with extra (Table 4)

For the CAS outcome in our adjusted models, we observed a statistically significant association between loss of job or wages because of COVID and CAS scores in the model adjusted for AAHLS total score (Model 1). Participants who reported that they lost their job or wages, had a statistically significant 1.30-point higher CAS score compared to participants who reported that they did not lose their job or wages (β=1.30, 95% CI [0.07, 2.53]). This estimate was fairly consistent in Model 2 (adjusted for AAHL and education) and model 3 (adjusted for AAHLS and financial situation at home), with estimates 1.30 (β=1.30, 95% CI [0.08, 2.52])) and 1.28 (β=1.28, 95% CI [-0.00, 2.55]) respectively (Table 4).

For the OCS outcome, we observed a statistically significant association between exercise during COVID and OCS scores in the model adjusted for AAHLS total score (Model 1). Participants who reported that they exercised during COVID, had a statistically significant 1.08-point higher OCS score compared to participants who reported that they did not exercise (β=1.08, 95% CI [-0.01, 2.16], p=0.05) In the model adjusting for AAHLS total score and education (model 2) and the model adjusting for AAHLS and financial situation (model 3), there were no statistically significant associations (Table 5).

### Emergent Themes from Thematic Analysis

Using the framework of social networks and social support, there were three domains of interest: pandemic impacts (Table 6), COVID-19 adaptations (Table 7), and Social Support Networks (Table 8). See Figure 2 for a summary of the themes within the context of the conceptual model. Emergent themes are further outlined below.

**Figure 2:**
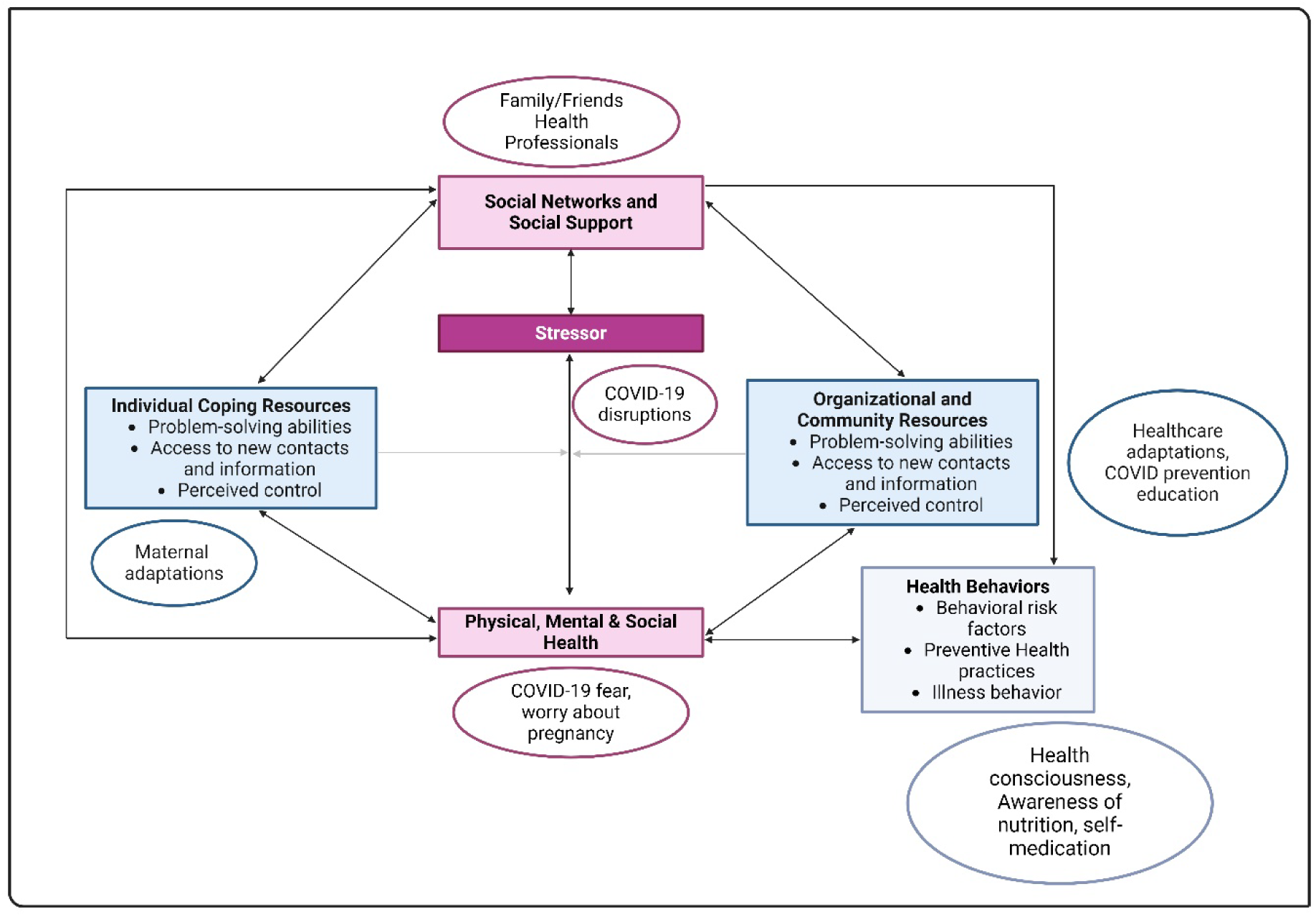
Analysis of the Impacts of the COVID-19 Pandemic within the Framework of Social Networks and Social Support to Health. Framework adapted from Heaney & Israel (2008).^41^ Themes generated from qualitative analysis are shown in the oval shapes. Created with BioRender.com.

**Table 6:** Themes from Qualitative Analysis: COVID Pandemic Impacts Domain.

| Sub-Domain | Themes | Example Quotes |
| --- | --- | --- |
| COVID-19 Disruptions | – Disruptions to healthcare delivery<br>– Disruptions to activities of daily living<br>– Disruptions to livelihood | ***“I wasn’t able to go to the clinic as expected every month. I stayed home for almost three to five months before going. When they said things were moderate you can go...I was...scared because, going, the doctors... have to touch your stomach...”***<br>***“I was not so worried; just that you couldn’t frequently go to the hospital, so you just had to be extra careful with the things you eat and then your movements. And wherever you’re going, just be careful you don’t get in touch with a lot of people. So... I was restricted, in the house most of the time... the only challenge I had was not to get Covid-19.”***<br>***“I didn’t miss the schedules. Just that at times when you’re not feeling so well, under normal circumstances you’ll...just go and check what is going on. But in this instance, when [I’m] not so well, [I] try to...wait until it’s the day for me to go for my appointment. Then when I go, I’ll mention all the issues. So that was what was happening. So, instead of under normal circumstances [when I] just go and come back before the appointment day, here... I was...holding on until the appointment day before I could go and explain whatever I was experiencing.”***<br>***“Even to go out and buy foodstuff and come and cook for the family was a problem. Because you don’t know who has it—either the person you’re going to buy the things from is having... Because they say it’s transferable—so even going out was very, very hard.”***<br>***“...before the Covid I can go [to the market] twice in a month or even thrice in a month. But because of the Covid, the advice of not exposing yourself to a lot of crowd... you might not know who’s having it; so, I’ll just try my best to do the shopping at once.”*** |
| COVID-19 psychosocial impacts | – Fear of catching COVID<br>– Worry about pregnancy<br>– Disruptions to social support networks | ***“...when it [the pandemic] came, at the beginning... how people were dying in China and others; how they were saying the spreading and other things, I was very scared.”***<br>***“...the pandemic brought us so much distress...because we were seeking for help [and] assistance and now there's Covid, we were all afraid as in, would we get the required help assistance we needed? Or what is going to happen to our babies? We had these thoughts in mind.”*** |
|  |  | “...my husband...told me at his workplace people were getting it [infection with the virus]; so that time **we all got worried**. He told me that some people had already contracted it, but they are all going to do the screening. So...they organized a screening for all of them...but he didn't get it.”<br>“...if it's not during the Covid, maybe I'll have to travel to the village with the kid. ...if your time is up, [and] you have to wait and deliver here—you have to do everything by yourself. But if [I] should have travelled to the village, [I'll] get hands to help, [I'll] get helpers around [me].”<br>“My sister was a health professional so I... **had to take extra precaution**, particularly with her. So, when she comes to the house, we make sure she has washed her hands, she's removed her clothes, everything, before she comes close to the children...she was actually at the emergency. So, she'll go and when she comes back, we're all like okay, for two days, you stay here; we'll try and leave you alone. It was quite an experience, but **we were hoping and praying none of us gets it**. And we were able to sail through...” |
| COVID-19 Opportunities | – Health consciousness<br>– Awareness of nutrition<br>– Time away from work | “I think the only thing was Vitamin C. I remember that one, so **I was taking Vitamin C**... the tablets and dissolve it and drink. That's what I was doing. But aside my eating was okay because I could eat well. **I was managing with fruits, and then vegetables.**”<br>“I adopted [the eating for two] style, since it was my first pregnancy...**I made sure my nutrition contained all the good nutrients; all the good foods**. And secondly, I realized that not all the cravings must be adhered to, like, because sometimes I get the cravings, but I don't even eat a handful of it.”<br>“[Before COVID] I was active. I could wake up in the morning, prepare the kids for school then also go to work. Then we'll come in the evening, let them do their homework, prepare food, eat, and then we'll go to sleep. So, **whiles we were home, once in a while, I'll take them walking in the evening before I sleep**. I'll just tell them I'm going for a walk, so let's walk because we've been in the house. So, we'll just stroll around this area for some time...” |
|  |  | *“... because I'm a teacher and then during that time, there was lockdown, so **for almost a year schools were closed so we weren't going to school.** So, I was just in the house.”* |

**Table 7:** Themes from Qualitative Analysis: COVID-19 Pandemic Adaptations.

| Sub-domain | Themes | Example Quotes |
| --- | --- | --- |
| Healthcare delivery adaptations | – Limits on hospital appointments<br>– Safety measures during hospital visits | *“When I was pregnant, by then the hospital I was attending my ANT [antenatal], they were attending to only emergency cases... so **[if we] were not having any complications in our pregnancy, we were asked to be in the house** because there was an outbreak.”*<br>*“Since the Covid started, **we couldn’t go for weighing**...unless you have to go for this immunization, they’ll advise that don’t come with... don’t bring the child. So, if you are coming for just weighing, they’ll tell you no. But if you’re coming for maybe polio or those things then you bring the child. So, for some months we were not going. I think we missed like three, four months.”*<br>*“At first when you go [for antenatal], you lie on their bed; **but due to the Covid, when you’re going you have to take your own rubber [plastic sheets/covers]**. Instead of lying on the bed, you have to lay your rubber on the bed and then you lie on it. So that you fold your thing and then put it in your bag [afterwards]. So, if you’re going and you don’t have, you have to buy. Due to Covid, they introduced that. Till now it’s still working.”* |
| Maternal Adaptations | – Self-limitation on hospital visits/change of providers<br>– Information seeking<br>– Self-medication<br>– Self-efficacy | *“...when it [COVID-19] came... they were announcing that if emergency for hospital or something like that you can go but me, I was scared. We are easily attracted to diseases and sicknesses due to our nature when you’re pregnant... So, I just took the book of one of my child’s and I saw the drugs... It’s repeating anytime you go—that’s the folic acid, multivitamins, and some blood tonic. And moreover, I was using abedru [turkey berries] as my blood tonic in the house—blending it and then drinking. And taking the folic acid. That was what I was doing till it was... it was somehow coming down then I started going to the hospital.”*<br>*“It got to a time that I could not feel my baby kick. So, I decided to go there [main government hospital] hoping that probably I would get the service of ultrasound. But I didn’t get it there, so **I had to go to a private hospital because I was anxious**. So, when I now got to the private ultrasound, they told me that my baby had an anterior placenta making me not to hear the fetal kicks, but my baby is fine. It was from there that I was somehow relaxed because it was even getting to 24, 26 weeks, and I was not feeling it* |
|  |  | *that much like how I used to feel my first pregnancy. I was not feeling it, so I was actually anxious.”* |
| Public COVID-19 education | – Prevention strategies<br>– Actions to take when symptoms arise | *“...avoid contact, wash your hands frequently, make sure you’re wearing your nose mask and... follow all the safety precautions... If you get Covid, call the hospital first before you come; if you’re experiencing some of the symptoms, call them before you come. These were some of the things [information] that we received.”* |

**Table 8:** Themes from Qualitative Analysis: Social Support Networks.

| Sub-domain | Themes | Example Quotes |
| --- | --- | --- |
| Sources of Social Support | – Family/friends<br>– Healthcare workers<br>– Paid labor | ***“I got an assistant [helping with shopping] I can't be heavily pregnant and still go to the market, no! I was treated like a [queen] during this time.”***<br>*“...[after delivery] a friend of mine, a colleague too just brought mashed kenkey for me in the hospital. And then in the afternoon two other colleagues at work also came. They ... brought me... fresh groundnuts...And then in the evening I came home with my husband.”*<br>***“Family people came, old ladies came and the siblings—not my children per se, but it's a family house so they all wanted to see the baby.”*** |
| Types/nature of Social Support | – Help with house chores<br>– Help with infant care | ***“So, along the line my sister visited me. She also stayed for about a week and then she was preparing food for me....In the mornings, she'll call me to go and eat. And she was also bathing the baby.”***<br>*“Before Covid I was the one shopping, but at times too he'll [her husband] go because of my daughter. At times I'll leave her with him and go, or he'll just go and buy the things. But most of the times, I was the one going to do the shopping. But when Covid-19 came, and at that time I was pregnant as well, we decided that...to protect all of us, I'll be in the house and then he'll do the shopping.”* |
| Impact of Social Support | – Help understanding health recommendations<br>– Advice on infant care<br>– Time for rest | *“...when you go and tell the doctors that my daughter is not eating, they're like: “what are you feeding her? how often?” Then I say I'm following the book—the red book. And at times I'm so confused; it's like I'm not doing something right, I don't know...they told me to add groundnut paste to the porridge. How will I add groundnut paste? I don't even know how... honestly, I think it's recently I got to know how to add it. I came back, I'll prepare the porridge, when the porridge is ready and I'm about to feed her, that's when I add the groundnut paste to it. I didn't know, so it was later my sister was like no, when you're preparing it, you can add it so that it will also get well cooked, then you add some powdered fish to it. It will help her.... And she was saying that don't feed her like what is in the book. So every two hours, try to give her something to eat. Even in the night, when she's sleeping, wake her up, let her eat something. So that was what I was doing—I was doing like round the clock feeding.”* |

#### COVID Pandemic Impacts

The COVID-19 pandemic impacts involved information pertaining to physical, mental, and social stressors associated with the pandemic, as well as behavioral risk factors and preventative health behaviors. Three emerging themes were used to divide this domain into subdomains: COVID-19 Disruptions, COVID-19 psychosocial impacts, and COVID-19 opportunities.

The emergent themes for the COVID-19 disruptions domain were disruptions to healthcare delivery, disruptions to activities of daily living, and disruptions to livelihood. As outlined in Table 6, while some participants shared that they missed scheduled hospital appointments, others shared that they restricted their hospital visits to only the scheduled ones and sometimes did not go to the hospital in between visits even when they had problems. In addition to the disruptions in health care delivery systems, participants also shed light on how the pandemic impacted their daily activities, including their ability to go shopping, and impacts on sources of livelihoods for those who were engaged in informal work.

The main psychosocial impacts identified were fear of catching COVID, worry about pregnancy, and disruptions to social support networks. Restrictions of visitors at the hospital post-delivery impacted social support during this time. Upon hospital discharge, some mothers also held off visits for several weeks, while others postponed travel to their village where they report that they could have obtained help from relatives (Table 6).

The emergent themes for the COVID-19 opportunities were health consciousness, awareness of nutrition, and time away from work. Due to the understanding that nutrition played a vital role in health, some participants were interested in finding health information that they could use to “boost their immune system.” Others adopted healthy lifestyles overall. Those who did not have to go to work due to the lockdowns also mentioned that this was positive for them. For example, a participant who was a teacher mentioned that she did not lose her wages when she had to stay home, and so this was helpful (Table 6).

#### COVID Pandemic Adaptations

For the domain of COVID-19 adaptation (Table 7), interest was in coping with the pandemic at both the individual and community levels. The themes included healthcare delivery adaptation, maternal adaptation, and public COVID-19 education. From the interviews, it appears that many public and government hospitals, especially those that served as national hubs for testing and isolating COVID-19 patients, adopted a triage system in which pregnant mothers and mothers with newborns were discouraged from visiting the hospital outside of their scheduled antenatal or immunization visits unless it was an emergency. The hospitals also incorporated safety measures into their practice to prevent the spread of the virus. For example, a mother discussed how hospitals required mothers to provide plastic sheets for covering their beds during antenatal exams (Table 7). Mothers also made personal decisions to limit visits to major hospitals for their own safety and adapted to the situation in diverse ways, including seeking information from online sources, self-medication, and self-efficacy in problem solving, especially for mothers who had had multiple children. Some mothers also switched their healthcare to private facilities and stopped utilizing the public facilities all together. All mothers interviewed mentioned that they had exposure to education on COVID-19 preventive measures.

#### Social Support Networks

The Social Support Networks domain (Table 8) involved information about the nature of social support received by mothers and how these helped mitigate stress during the postpartum period. Emergent themes for this domain included sources of social support, types of social support, and impact of social support. Even though some mothers reported disruptions to social support systems as mentioned above, most participants interviewed reported that they had some form of social support system during pregnancy and when they delivered their baby. The main sources of social support were family, friends, and healthcare workers. Some of the strongest ties were participant’s partners, siblings (sisters and sisters-in law), and parents (mothers and mothers-in law). Some of the weaker ties included friends and health professionals during the antenatal care and delivery process. The nature of support included help with chores and help with infant care (including help with other children besides the index child). The main impacts of social support were adherence to and/or help understanding health recommendations, advice on infant feeding, and more time to rest due to time away from chores.

## Discussion

In our study, we first set out to retrospectively determine the prevalence of COVID-related anxiety during pregnancy among Ghanaian women using an online and phone survey. We then assessed the relationship between COVID-related impacts on COVID-related anxiety and obsession. To contextualize our findings and understand the impact of social support on maternal COVID experiences, we also engaged with participants to collect qualitative information regarding their experiences of pregnancy and childcare during the pandemic.

We observed a rate greater than 10% for probable dysfunctional coronavirus-related anxiety and even higher for probable dysfunctional thinking about COVID-19 in our study population based on the coronavirus anxiety scale and obsession with coronavirus scale (11% and 33%, respectively). Because this, to the best of our knowledge, is the first study to use the CAS and OCS scales among Ghanaian women, there is no baseline COVID-19 anxiety rate to compare this to. One study we found conducted among 71 Ghanaian women assessed COVID anxiety using two questions related to being worried/thinking too much about being pregnant during COVID and being very worried/thinking too much about giving birth during COVID-19.^23^ They found that 66.2% of participants were thinking too much about being pregnant during COVID-19 and 71% of their participants were thinking too much about giving birth during COVID-19. Because excessive thinking about COVID is closest to our measures of obsession, these are most comparable to our OCS measures, suggesting that dysfunctional thinking about the pandemic was widespread among the population of pregnant women during the pandemic. We did not find any other studies based in Ghana examining COVID-specific anxiety, highlighting the strong need for more specific, standardized tools tailored to the African context in future pandemic situations.

In a non-Ghanaian study that specifically used the CAS scale among Indian women infected with the coronavirus, about 18.3% of antenatal mothers and 14.6% of postnatal mothers were suffering from dysfunctional anxiety,^37^ which is higher than the 11% that we observed in our sample using the CAS scale. It is important to note that our participants had already delivered their babies at the time they were completing our survey and had to recall their prior experiences during the pandemic. As observed in the study by Najam and colleagues, more women seemed to have COVID-related anxiety during pregnancy compared to postpartum women.^37^ Additionally, the reported COVID-19 infection rate in our population was almost negligible (2%) as opposed to their cohort, who were all infected with COVID. It is therefore possible that our participants’ recollection of their pregnancy-related anxiety was biased by their postpartum experiences, potentially causing a shift in perception of COVID-19 risk, coupled with the fact that the reported caseloads for COVID were lower in Ghana than in other countries by 2021.

A study among Turkish pregnant women observed mean CAS and OCS scores of 5.38 ± 4.55 and 2.98 ± 3.12 respectively.^47^ Their mean CAS scores were higher than those we observed (3.5±3.5), while their OCS scores were lower than ours (5.4±3.6). A similar Turkish study that used both the CAS and OCS scales among hospitalized high-risk pregnant women observed mean rates of 0.7 ± 1.9 and 1.7 ± 2.3, respectively,^38^ which were far lower than the rates we observed in our cohort as well as the earlier rates observed in the same country. Both these studies did not report the proportion of their participants who met the criteria for dysfunctional CAS and OCS scores, however, the fact that their results were vastly different warrant discussions on the relevance of timing for assessing pandemic related anxiety. The earlier study assessed participants between November 2020 and June 2021 whereas the latter study assessed participants between March 2021 and March 2022, which may suggest that dysfunctional anxiety might have peaked during the early stages of the pandemic. Our study recruited participants between June 2021 and January 2022 and asked them to retroactively recall their COVID experiences. While we asked participants to recall events of their pregnancies, which would have been between 2020 and 2021, our time frame potentially impacted recall. Nevertheless, for a country like Ghana that did not have as high a reported case load as Turkey as of 2021,^48^ our observed rates of COVID-19 anxiety and obsession warrant safeguards for additional psychosocial support during future pandemics.

A source of COVID-related anxiety we found in our qualitative analysis was the fear of catching COVID, as well as concerns about pregnancy and the safety of the unborn child, which are in keeping with studies conducted by Moyer et al. (2021) in Ghana, Akgor et al. (2021) in Turkey, and a literature review by Adu et al (2022).^23,49,50^ These results are also in agreement with a recently published qualitative study that observed fears among pregnant women amidst the pandemic,^51^ as well as studies linking fear of COVID-19 with COVID-19 anxiety.^14,52^ COVID-19 fear could also be the explanation for the high prevalence of probable dysfunctional thinking about coronavirus (33%) that we observed in our population. The intense fear of contracting an infectious disease that saw extensive media coverage seems rational and expected, especially given that this was a novel virus, leading to dysfunctional thinking about the pandemic. This fear could also explain the observed statistically significant positive association between exercise and high OCS scores. Our qualitative data suggests that participants adopted health-conscious behaviors during the pandemic to reduce their perceived risk of catching the virus. It is therefore possible that exercise during the pandemic predicted high OCS scores due to health vigilance driven by the fear of illness.

In all our predictive models, we also found that job or wage loss significantly increased coronavirus anxiety, with an average rise of 1.30 points on the Coronavirus Anxiety Scale. This outcome is unsurprising, as losing a source of income introduces substantial stress and uncertainty, which can intensify the psychological impact of the pandemic. However, because CAS is a relatively new tool—especially within African populations and among postpartum women—further research is needed to fully understand the clinical implications of a 1.30-point increase. Notably, CAS uses a 0–20 scale, with a score of 9 or above indicating probable dysfunctional coronavirus-related anxiety. This suggests that income loss may have a particularly strong effect on individuals who are near this threshold (those scoring around 7 or 8), potentially pushing them into the clinically significant range. Consequently, public health strategies aiming to identify individuals at risk of pandemic-related psychological distress might consider adjusting the CAS threshold for at-risk classification and integrating economic support mechanisms to mitigate the mental health impact of lost livelihoods.

It is also crucial to discuss our observed lack of association between other COVID-related socioeconomic impacts (financial situation) and coronavirus anxiety and obsession in our population, which could be attributed to the presence of social support systems in our population. A study among women of color in the US found that COVID-19 significantly increased levels of COVID stress, which significantly increased rates of depression and reduced resilience among study participants.^53^ Some of the factors contributing to stress in their study included loss of income and COVID-19 Isolation, which we observed in both our quantitative data and qualitative interviews. In our study, however, we found that social support was readily available to our participants, even among those who decided to isolate.

Of note, Ghana has a communal culture where pregnant and postpartum women typically receive emotional and instrumental support from their social networks, including well-being checks, advice and guidance, help with household chores and food, and even monetary assistance before, during, and after delivery.^54,55^ These collectivist norms ensure that vulnerable members of society, and especially pregnant women, cope with daily stressors. Additionally, low social support has been shown to mediate high maternal mental health disorders among Ghanaian women.^56^ We hypothesize that the abundant reports of social support among our cohort may have resulted in better cognitive appraisal and coping strategies, which have been shown to impact COVID-related mental health outcomes in other cohorts.^53,57–59^

While plausible, this may, however, not be the only explanation for our observations. We observed a trending relationship between financial hardship (that is, struggling to make ends meet) and coronavirus anxiety in our unadjusted model, which was attenuated when we adjusted for other maternal characteristics. On the reverse, while there were no associations between financial hardship and coronavirus obsession in our unadjusted models, we began to see a trend between financial hardship and COVID obsession in adjusted models, although this did not reach statistical significance. The financial hardship measure we used was a 4-point ordinal measure with only a few of our participants on either side (those who were doing extremely well with extra income versus those who were struggling to make ends meet), likely making it a less sensitive measure compared to the actual loss of wages described previously. Majority of respondents reported that they were just doing ok or had to cut back. We therefore cannot rule out a lack of statistical power to detect true differences if indeed there were any. Additionally, our weak associations may suggest the presence of some residual confounding, or that there could have been latent or unobserved factors mitigating the relationship between financial hardship and COVID-related psychosocial health that were not measured in our study. Finally, the fact that financial hardship had opposite effects on anxiety versus obsession warrants additional inquiry and might suggest unique pathways by which stressors impact psychological health. For instance, because coronavirus anxiety assesses psychosomatic symptoms while coronavirus obsession is concerned with disturbed thinking about the pandemic, the latter being more widespread in our population compared to the former, our predictors may have different effects on the relationship if they have varying abilities to trigger physical reactions to stress compared to thinking or worry.

Of importance, we also observed that higher health literacy (either functional, critical, communicative, or all aspects combined) was significantly associated with lower scores on our anxiety scale while controlling for other maternal characteristics like age, educational status, and parity. This speaks volumes about the important and often understudied relationship between health literacy and mental health, warranting more investigation into this relationship. If health literacy and other unmeasured characteristics were potentially mediating the response between the socio-economic impacts of the pandemic and the psychosocial well-being of our participants, future studies exploring long-term pandemic-related mental health could consider assessing latent socio-economic patterns to provide a more holistic picture of the socio-economic dynamics that predispose vulnerable women to pandemic-related mental health impacts.

## Strengths and Limitations

Our study had many strengths. First, because it was conducted online during the time of the pandemic, it allowed study investigators to collect a wide range of information regarding Ghanaian maternal experiences of the COVID-19 pandemic, without the need for physical contact. This came with the unintended consequence of sampling mostly women who had access to the internet. We tried to mitigate this by including phone calls to mothers who did not have access to the internet or were not computer-literate enough to self-administer the survey. Despite these efforts, our study was heavily weighted toward women who were highly educated. Indeed, over 60% of our participants had a tertiary education or higher. Additionally, because this study was conducted online and over the phone, our study sample appears to be skewed towards potentially higher-income, highly literate women, whose COVID-19 experiences may differ from those of low-literacy or lower-income women. While we made efforts to station research assistants in the northern, middle, and southern belts of the country, our study sample was not a nationally representative sample, and therefore, our findings cannot be generalized. Another key strength of our study is the mixed methods approach we employed. Incorporating the voices of our participants, particularly regarding their experiences with social support, into the interpretation of our study findings enabled us to derive more meaning from the survey results than we would have otherwise.

A limitation we had was our inability to make comparisons between pre-and post-pandemic anxiety rates, a consequence of our novel use of the CAS and OCS measures in the Ghanaian population. The CAS instrument used in our study is only meant to serve as a screening tool for COVID-related stress and not as a diagnostic tool for anxiety, stress, or other depressive conditions. For this reason, we are unable to make a direct comparison between the rates of stress and anxiety pre– and during the pandemic, given that the data we had on pre-pandemic depressive rates were based on instruments like the Edinburgh postpartum depressive scale (EPDS) and the patient health questionnaire (PHQ). Nevertheless, we chose the CAS and OCS tools because of their specificity regarding the COVID-19 pandemic. Both the PHQ and the EDPS might be useful in diagnosing antenatal and postpartum depression but are not designed to specifically capture pandemic-related stress, which was the focus of our research. Additionally, as we discussed earlier, our study is not nationally representative or generalizable, and relied on self-report and recall, which are prone to recall bias. If participant’s recollection of their pregnancy-related anxiety experiences were influenced by their postpartum experiences, then it is likely that our estimates are an underestimate of the true COVID-19 related anxiety rates.

## Implications for Research and Practice

The world has seen pandemics before, and the COVID-19 pandemic is unlikely to be the last one of this scale. However, the pandemic can be a historical relic from which we learn lessons for future resilience. Globally, rates of anxiety and depression stemming from fear of contracting the virus, and disruptions to day-to-day activities were high, as reflected in our data, which may warrant the integration of psychosocial screening into antenatal care for pregnant women especially during catastrophes. This also raises the question: In what ways can we mitigate the psychosocial insults caused by an infectious disease pandemic necessitating restrictions on movement and physical contact? One answer lies in our ability to leverage modern technologies that help us stay connected with each other and with primary health providers. Not only will this work to extend social support networks but also provide platforms for mobile maternal mental health support. Further, as Davenport and colleagues noted, and corroborated by our findings, the pandemic reduced maternal attendance to health care visits necessary for early psychological diagnosis and treatment of depressive symptoms; therefore, employing remote strategies for reaching women during periods like these is necessary.^60^

One of the most interesting findings of this study is the general disconnect between certain COVID-related disruptions and COVID-19 related stress and anxiety. These likely were mitigated by the abundance of social support in these contexts and the public health education that participants reported receiving. Indeed, from our qualitative analysis, it appears that a desire to modify lifestyle behaviors for the better was a direct consequence of these stresses related to the pandemic. Because the sample size of our project is small, and not nationally representative, this does raise questions about whether experiences were different for other populations of women, specifically those who resided in more impoverished and resource limited settings and unable to afford private facilities when the public institutions got overwhelmed.

Future studies should also iteratively engage additional theoretical models of human behavior. While the main framework for the formulation of our research questions was informed by the Transactional Model of Stress and Coping, it was imperative to engage the Social Network Theory in understanding our findings, especially those regarding the role of social support in maternal and infant health outcomes. In our sample, we found that most of our participants, despite the COVID-19 disruptions, had social support which might have mitigated the negative impacts of the pandemic. This is key to understanding the possible reasons why, despite a global catastrophe, mothers in this part of the world were able to adapt to their stressful birthing and postpartum experiences.

Another answer lies in strategies to improve maternal health literacy. For future studies, health interventions in this population must leverage the social networks of mothers and children as key players in 1) disseminating health information, 2) implementing health education recommendations at the family and community levels, and 3) monitoring and evaluating health education interventions. Studies also integrating other dimensions of health literacy, specifically as they relate to maternal health literacy, are also warranted. Finally, policies integrating psychosocial screening into the antenatal care framework as well as those strengthening mobile and telehealth systems should be reinforced.

## Declarations

### Ethics approval and consent to participate

Institutional Review Board (IRB) approval in accordance with the principles outlined in the Belmont Report was obtained from the University of Massachusetts Amherst (IRB: #2578). Additionally, the study adhered to the guiding principles of the Declaration of Helsinki. All participants completed and signed a consent form prior to enrollment in the study. Participants who could not access the project online provided verbal consent after interviewers explained the details of the project as outlined on the consent form to them.

## Availability of data and materials

The datasets used and/or analyzed during the current study are available from the corresponding author on reasonable request.

## Authors’ contributions

R.A conceptualized and designed the study under the guidance of EC, GO, JP and SGN. RA managed the data in REDCap and performed the data analysis in SAS. All authors contributed to the interpretation of the data. RA wrote the first draft of the manuscript. All authors read, critically reviewed, and contributed to substantive revisions of the first draft. All authors agreed on the final manuscript.

## Data Availability

The datasets used and/or analyzed during the current study are available from the corresponding author upon reasonable request.

## Acknowledgements

RA would like to extend her appreciation to the UMASS Graduate School and the MMEG grant for their financial support. To the research assistants in Ghana who helped with data collection and data quality control for phase 1 of the study, Matilda Amanor, Ohui MacCarthy, and Hisham Osumanu, the authors would like to say a very big thank you!

