## Supplemental data for "A Retrospective Mixed Methods Analysis of COVID-19’s Impact on Maternal Psychosocial Health in Ghana: Lessons for Future Public Health Crises"

### Supplemental Information 1: PREPARE Screening and Eligibility form

PREPARE PROJECT_ACTIVE PID 7559

Actions: Download PDF of instrument(s) Share instrument in the Library VIDEO: Basic data entry

#### PREPARE Screening, Eligibility form

Adding new Participant ID **1222**

Event Name: **Baseline**

**Participant ID** 1222

**Name of Interviewer (for interviewer administered questionnaires)**

**Date of interview (for interviewer administered questionnaires)** Today D-M-Y

**Contact Details**

**Please tell us your name**

**First Name**

**Last Name**

**Area of Residence (Specify)**
*This is the area where you live. Eg. Accra, Kumasi, Tamale etc*
* must provide value

**Region (please select your Region from the dropdown list)**
* must provide value

**Regional zone** Southern View equation

**Coded regional zone**
- Northern
- Southern

reset

**Phone number**
* must provide value

**Email address**
*The project will use this email address to communicate with you and remind you about questionniares. It is important that you continue to check this email for follow-up questionnaires.*
* must provide value

**Maternal Eligibility**

**How old are you? (in years)**
*Provide your age in years*
* must provide value

**Are you currently pregnant?**
* must provide value
- Yes
- No

reset

**Please let us know if you currently have a child**

Do you have a child between 0-15 months?

**Note:** Mothers completing in November 2021 can have a 15 month old child. **Please select yes if you have a child up to 15 months old**
* must provide value
- Yes
- No

reset

**Were you diagnosed with depression during or after your pregnancy?**
* must provide value
- Yes
- No

reset

**Do you have any special condition we should know of?**
* must provide value
- Yes
- No

reset

**Details for Mobile Money transfers**

| | |
|---|---|
| **Exclude from baseline phase 1?** | ○ Yes<br>○ No<br>reset |
| **Reason for phase 1 exclusion** | ☐ Blank record<br>☐ Duplicate record<br>☐ Incomplete data<br>☐ Automatic (ineligible)<br>☐ Invalid (based on Matilda's DQC)<br>☐ Other |
| **Form Status** | |
| **Complete?** | Incomplete |
| **Lock this instrument?**<br>If locked, no user will be able to modify this instrument for this record until someone with Instrument Level Lock/Unlock privileges unlocks it. | ☐ Lock |
| | Save & Exit Form Save & Stay<br>-- Cancel -- |

**Supplemental Information 2: Baseline Demographic Survey**

PREPARE PROJECT_ACTIVE PID 7559

Actions: Download PDF of instrument(s) | Share instrument in the Library | VIDEO: Basic data entry

#### Baseline Demographic Survey

Adding new Participant ID **1222**

Event Name: **Baseline**

| | |
|---|---|
| **Participant ID** | 1222 |
| **Section 1: Background and Socio-Demographic Data** | |
| **Is this your first child?**<br>* must provide value | ○ Yes<br>○ No<br>reset |
| **Have you ever visited a dietitian or nutritionist for dietary counselling of any sort?**<br>* must provide value | ○ Yes<br>○ No<br>reset |
| **Marital Status**<br>* must provide value | ○ Married (this includes customary/"traditional", "church" and civil/court marriage)<br>○ Living with partner but not married (Cohabiting)<br>○ Single<br>○ Divorced<br>○ Widowed<br>reset |
| What is your **completed** level of education? (this is the highest level of education that you have completed)<br>* must provide value | ○ No school<br>○ Basic school (up to JSS 3)<br>○ Secondary (SHS/Vocational/Technical)<br>○ Tertiary (University or Training College)<br>○ Postgraduate (Masters, PhD or other post baccalaureate degree)<br>reset |
| **What job do you do? (What's your occupation?)**<br>* must provide value | ○ Unemployed<br>○ Public institution<br>○ Private company<br>○ Business/market woman/petty trader<br>○ Housewife<br>○ Other<br>reset |
| **What is your estimated monthly income?**<br>* must provide value | ○ Less than GHC 500<br>○ GHC 500-999<br>○ GHC 1000-1999<br>○ GHC 2000-3000<br>○ More than GHC 3000<br>reset |
| **What is the average monthly family expenditure (on food and utilities only)?**<br>*This is how much money the whole family spends on food, electricity bills, water bills etc.*<br>* must provide value | ○ Less than GHC 200<br>○ GHC 200-499<br>○ GHC 500-699<br>○ GHC 700-1000<br>○ More than GHC 1000<br>○ Don't know<br>reset |
| **Form Status** | |
| **Complete?** | Incomplete |
| **Lock this instrument?**<br>If locked, no user will be able to modify this instrument for this record until someone with Instrument Level Lock/Unlock privileges unlocks it. | ☐ Lock |
| | Save & Exit Form · Save & Stay<br>-- Cancel -- |

https://arcsapps.umassmed.edu/redcap/redcap_v11.0.1/DataEntry/index.php?pid=7559&page=baseline_demographic_survey&id=1222&event_id=19…

#### Supplemental Information 3: All Aspect of Health Literacy Scale Questionnaire

**PREPARE PROJECT_ACTIVE** PID 7559

Actions: Download PDF of instrument(s) | Share instrument in the Library | VIDEO: Basic data entry

##### All Aspect of Health Literacy Scale

Adding new Participant ID **1222**

Event Name: **Baseline**

**Participant ID** 1222

**Health Literacy**

**Please tick one response only for each question by ticking the radio button.**
If you'd prefer, a staff member of the research team can read out questions and explain them to you. Email us at *or send a Whatsapp message to +233202502729* if you feel that you will need extra assistance completing the questionnaire.

Thank you!

**Functional Health Literacy**

| | Often | Sometimes | Rarely |
|---|---|---|---|
| **How often do you need someone to help you when you are given information to read by your doctor, nurse or pharmacist?** | ○ | ○ | ○ reset |
| **Do you need help to fill in official documents?** | ○ | ○ | ○ reset |

**Communicative Health Literacy**

| | Often | Sometimes | Rarely |
|---|---|---|---|
| **When you talk to a doctor or nurse, do you give them all the information they need to help you?** | ○ | ○ | ○ reset |
| **When you talk to a doctor or nurse, do you ask the questions you need to ask?** | ○ | ○ | ○ reset |
| **When you talk to a doctor or nurse, do you make sure they explain anything that you do not understand?** | ○ | ○ | ○ reset |

**Critical Health Literacy**

| | Often | Sometimes | Rarely |
|---|---|---|---|
| **Are you someone who likes to find out lots of different information about your health?** | ○ | ○ | ○ reset |
| **How often do you think carefully about whether health information makes sense in your particular situation?** | ○ | ○ | ○ reset |
| **How often do you try to work out whether information about your health can be trusted?** | ○ | ○ | ○ reset |

**Are you the sort of person who might question your doctor or nurse's advice based on your own research?** ○ Yes, definitely ○ Maybe/sometimes ○ Not Really reset

| Empowerment | |
|---|---|
| **Do you think that there are plenty of ways to have a say in what the government does about health?** | ○ Yes, definitely ○ Maybe/sometimes ○ Not Really<br>reset |
| **Within the last 12 months have you taken action to do something about a health issue that affects your family or community** | ○ Yes ○ No<br>reset |
| **What do you think matters most for everyone's health?**<br>You can choose **only one** answer | ○ a) information and encouragement to lead healthy lifestyles<br>○ b) good housing, education, decent jobs and good local facilities<br>reset |
| **Form Status** | |
| **Complete?** | Incomplete |
| **Lock this instrument?**<br>If locked, no user will be able to modify this instrument for this record until someone with Instrument Level Lock/Unlock privileges unlocks it. | ☐ Lock |
| | Save & Exit Form Save & Stay<br>-- Cancel -- |

**Supplemental Data 4: COVID-related Anxiety Scale (CAS)**

PREPARE PROJECT_ACTIVE PID 7559

Actions: Download PDF of instrument(s) | Share instrument in the Library | VIDEO: Basic data entry

#### COVID Related Stress

Adding new Participant ID **1222**

Event Name: **Baseline**

**Participant ID** 1222

**Coronavirus Anxiety Scale**

**How often have you experienced the following activities during your pregnancy?**

| | Not at all | Rare, less than a day or two | Several days | More than 7 days | Nearly every day for over 2 weeks in a stretch |
|---|---|---|---|---|---|
| **1. I felt dizzy, lightheaded, or faint, when I read or listened to news about the coronavirus.** | ○ | ○ | ○ | ○ | ○ reset |
| **2. I had trouble falling or staying asleep because I was thinking about the coronavirus.** | ○ | ○ | ○ | ○ | ○ reset |
| **3. I felt paralyzed or frozen when I thought about or was exposed to information about the coronavirus.** | ○ | ○ | ○ | ○ | ○ reset |
| **4. I lost interest in eating when I thought about or was exposed to information about the coronavirus.** | ○ | ○ | ○ | ○ | ○ reset |
| **5. I felt nauseous or had stomach problems when I thought about or was exposed to information about the coronavirus.** | ○ | ○ | ○ | ○ | ○ reset |

**CAS total score** [ ] View equation

**Form Status**

**Complete?** Incomplete

**Lock this instrument?**

If locked, no user will be able to modify this instrument for this record until someone with Instrument Level Lock/Unlock privileges unlocks it.

☐ Lock

Save & Exit Form | Save & Stay

-- Cancel --

#### Supplemental Information 5: Obsession with Coronavirus Scale (OCS)

**PREPARE PROJECT_ACTIVE** PID 7559

Actions: Download PDF of instrument(s) | Share instrument in the Library | VIDEO: Basic data entry

##### Obsession with Coronavirus

Adding new Participant ID **1222**

Event Name: **Baseline**

**Participant ID** 1222

How often have you experienced the following activities **during your pregnancy?**

| | Not at all | Rare, less than a day or two | Several days | More than 7 days | Nearly every day for over 2 weeks in a stretch |
|---|---|---|---|---|---|
| **1. I had disturbing thoughts that I may have caught the coronavirus.** | ○ | ○ | ○ | ○ | ○ reset |
| **2. I had disturbing thoughts that certain people I saw may have the coronavirus.** | ○ | ○ | ○ | ○ | ○ reset |
| **3. I could not stop thinking about the coronavirus.** | ○ | ○ | ○ | ○ | ○ reset |
| **4. I dreamed about the coronavirus.** | ○ | ○ | ○ | ○ | ○ reset |

**OCS total score** [ ] View equation

**Form Status**

**Complete?** Incomplete

**Lock this instrument?**

If locked, no user will be able to modify this instrument for this record until someone with Instrument Level Lock/Unlock privileges unlocks it.

☐ Lock

Save & Exit Form | Save & Stay

-- Cancel --

#### Supplemental Information 6: COVID-related Health and Wellbeing Questionnaire



**PREPARE PROJECT_ACTIVE** PID 7559

Actions: Download PDF of instrument(s) Share instrument in the Library VIDEO: Basic data entry

##### COVID-related Health and Well-being Questionnaire

Adding new Participant ID **1222**

Event Name: **Baseline**

| | | |
|---|---|---|
| **Participant ID** | 1222 | |
| **Lifestyle Changes**<br><br>This section will ask you some questions about any lifestyle changes you had during the pandemic. | | |
| **Are you currently living under a "lockdown" (shelter in place or shelter in home) order?** | ◯ Yes ◯ No ◯ I don't know | reset |
| **Did you ever live under a "lockdown" (shelter in place or shelter in home) order during your pregnancy** | ◯ Yes ◯ No | reset |
| **Do you personally know anyone who has had COVID? (this could be a friend or any member of your family)** | ◯ Yes ◯ No | reset |
| **Did you have COVID at any point during your pregnancy?** | ◯ Yes ◯ No ◯ I don't know | reset |
| **How likely do you think it is that you have COVID right now?** | ◯ Very unlikely<br>◯ Unlikely<br>◯ Somewhat likely<br>◯ Likely<br>◯ Very likely | reset |
| **Are you or anyone in your household currently self-isolating because of a suspected COVID-19 infection or exposure to someone with COVID.** | ◯ Yes ◯ No | reset |
| **Were you working in-person at (reporting physically to) your work site during your pregnancy** | ◯ Yes ◯ No | reset |
| **Did you lose your job or any salary (wages) because of the COVID-19 pregnancy?** | ◯ Yes ◯ No | reset |
| **Were you able to get any help at home (from friends or relatives)when you delivered?** | ◯ Yes ◯ No | reset |
| **How would you describe the money situation in your household right now?** | ◯ Comfortable, with extra<br>◯ Enough, but no extra<br>◯ Have to cut back<br>◯ Cannot make ends meet | reset |
| **What is the total number of people that live in your home?** (including yourself, all other adults, and children) **Note:** this refers to the number of people within your family who live together in the same house | ________ | |
| **During your pregnancy, did you work or volunteer in a hospital, emergency room, clinic, medical office, or nursing home, ambulance services, first responder services, or any healthcare setting or taking care of patients as a health professional, student or as part of your work?** | ◯ Yes ◯ No | reset |
| **This section of the survey will ask you about exercise**<br><br>Think back during the time that you were pregnant, and answer the questions that follow<br><br>*When you're ready to move on, click the submit button at the end of this section* | | |

**During your pregnancy, how many times a week on average did you do exercise for more than 10 minutes during your free time?**

- ○ No exercise (0 days)
- ○ 1 day
- ○ 2 days
- ○ 3 days
- ○ 4 days
- ○ 5 days
- ○ 6 days
- ○ 7 days (every day during the week)

reset

**Definitions** (for exercise intensity)

***Mild exercise***

e.g. easy walking for more than 10 minutes during your free time. (Note: if you engaged in not very common sports like yoga and golf, choose this category)

***Moderate exercise***

e.g. fast walking, dancing, easy bicycling, volleyball, tennis, easy swimming for more than 10 minutes during your free time

***Strenuous exercise***

e.g. running, jogging, football/soccer, basketball, vigorous long distance bicycling, vigorous swimming for more than 10 minutes during your free time

**All activity should have lasted more than 10 minutes to count as exercise.**

**Form Status**

**Complete?** Incomplete

**Lock this instrument?**

If locked, no user will be able to modify this instrument for this record until someone with Instrument Level Lock/Unlock privileges unlocks it.

☐ **Lock**

Save & Exit Form | Save & Stay

-- Cancel --

**Supplemental Information 7: Interview Guide**

**PREPARE Project:** Interviewer Guide-Phase 1

**Introduction**

Start interview by thanking woman for agreeing to participate and putting her at ease. Check to make sure she is still happy to be interviewed and to be audio/video taped. Describe the interview process and the need for honest, candid, and open answers (tell participant to answer as best as she can; that there are no right or wrong answers; and explain that you want their opinions and responses) and start the recording.

**Pregnancy and lactation**

- Tell me about your pregnancy experience during the coronavirus pandemic.
    - How worried were you about your pregnancy when you heard about the COVID-19 pandemic?
      **Note:** Probe for specifics, then follow-up with a 3-point scale: Very, somewhat, not at all
    - What were some of the worries you had related to COVID and pregnancy?
    - Did you receive any information related to COVID at the hospital?
- Were you able to attend all your **scheduled** antenatal sessions? How many were scheduled and how many were you able to attend out of them?
    - If not, why not?
- What challenges did you have accessing antenatal care during your pregnancy?
- Were you able to breastfeed after giving birth? [if yes, ask about onset of lactation e.g., when were you able to begin breastfeeding?]
    - During this period, was your child given anything to drink when you delivered? (Water, sugar water, formula etc.)-ask this question only if onset of lactation was not immediate.
- Were you able to breastfeed exclusively for 6 months? This means no water or other drinks or food for the first 6 months of your child's life. Why not?
    - Did the people at home provide enough support you to breastfeed? In what ways?
    - Did you feel pressured by family or close friends to give other things before 6 months? If yes, by whom, and what was their reason?
    - If you worked, did you feel like you had enough support at work to breastfeed?
- What challenges did you have accessing postpartum and infant care after you delivered?

**Covid-related health, social wellbeing, and financial impact.**

- How was your experience at the hospital during delivery (crosscheck from baseline survey if mother delivered at the hospital)

    - During your time in the hospital [if woman delivered in hospital], did you have any visitors? How about when you went home? Did you have any help at home after delivery? Who helped you?
- Were you working outside your home before the COVID-19 pandemic?
    - What job were you doing before the COVID-19 pandemic? Did this change?
    - How did the pandemic affect your employment?
- Tell me about your daily activities during your pregnancy (what are the different activities you did each day?)
    - What types of activities did you do during the pandemic?
    - Were you physically active before the pandemic? Did this change? How?
    - What impacted your ability to engage in physical activities? (If none)
- Were you able to go out to shop for food during this time? How many times did you have to go out to the market (to do grocery shopping)? [Was this an increase or reduction compared to pre-COVID times]

**COVID, health knowledge, and input on intervention**

- What do you wish you knew during your pregnancy? [what would you like to learn now about nutrition for yourself and for your child]
- Reflecting on the pandemic and how there was limited physical interaction due to COVID lockdown, how do you think health professionals can educate people? [in what ways can we provide information to others without physical contact]
